# Behavioral Inhibition and Dual Mechanisms of Anxiety Risk: Disentangling Neural Correlates of Proactive and Reactive Control

**DOI:** 10.1101/2020.12.01.20242123

**Authors:** Emilio A. Valadez, Sonya V. Troller-Renfree, George A. Buzzell, Heather A. Henderson, Andrea Chronis-Tuscano, Daniel S. Pine, Nathan A. Fox

## Abstract

**Background:** Behavioral inhibition (BI) is a temperament style characterized by heightened reactivity and negative affect in response to novel people and situations, and it is a strong predictor of anxiety problems later in life. However, not all BI children develop anxiety problems and mounting evidence suggests that how one manages their cognitive resources (cognitive control) influences anxiety risk. The present study tests whether more (proactive control) or less (reactive control) planful cognitive strategies moderate relations between BI and anxiety.

**Methods:** Participants included 144 adolescents (55.9% female) whose temperament was assessed during toddlerhood. In adolescence (*M*_*age*_ = 15.4 years), participants completed an AX Continuous Performance Test while EEG was recorded in order to disentangle neural activity related to proactive (cue-locked P3b) and reactive (probe-locked N2) control.

**Results:** BI was associated with greater total anxiety scores only among adolescents with smaller ΔP3bs and larger ΔN2s – a pattern consistent with decreased reliance on proactive strategies and increased reliance on reactive strategies. Additionally, a larger ΔP3b was associated with greater total anxiety scores.

**Conclusions:** BI relates to risk for anxiety specifically among adolescents who rely less on proactive strategies and more on reactive control strategies. Results further suggest that proactive control differentiates a BI-related etiological pathway to anxiety from a more general pathway to anxiety occurring regardless of BI level. Thus, developmental context (i.e., temperament) moderates the association between anxiety and proactive control. The present study is the first to characterize how proactive and reactive control uniquely relate to pathways toward anxiety risk.

---

Behavioral inhibition (BI) is a temperament style characterized by heightened reactivity and negative affect in response to novel people and situations (Kagan et al., 1984). BI is among the strongest predictors of later-life anxiety problems (Fox et al., 2005; Fox & Pine, 2012; Schwartz et al., 1999), especially when BI is stable throughout infancy and early childhood (Chronis-Tuscano et al., 2009). Nevertheless, 30-60% of toddlers with BI do not go on to meet criteria for an anxiety disorder during childhood or adolescence (Clauss & Blackford, 2012; Gladstone et al., 2005). Thus, identifying factors that moderate the relations between BI and anxiety remain a key issue for prevention and intervention.

Mounting evidence finds cognitive control, including use of specific cognitive control strategies, may be a marker of anxiety risk for children with BI (Henderson, 2010; Lamm et al., 2014; McDermott et al., 2009; Smith et al., 2019; Troller-Renfree et al., 2019). Dual-mechanisms of control theory (Braver, 2012) differentiates two temporally distinct and complementary, yet largely independent strategies of cognitive control: proactive and reactive. Proactive control involves early selection and maintenance of goal-relevant information, such as the biasing of attention toward stimulus color rather than the content of the word on the color-word Stroop task. Reactive control involves as-needed capacities, often in response to conflict, such as conflict between the color and word content on an incongruent Stroop stimulus, or conflict between the response that was made and the response that should have been made. During the first decade of life, children tend to shift from greater reliance on reactive control to greater reliance on proactive control (Lucenet & Blaye, 2014; Troller-Renfree et al., 2020). Afterwards, proactive control continues to grow in efficiency throughout adolescence and young adulthood (Chevalier et al., 2015).

A longitudinal study examining cognitive control factors in the context of BI-anxiety relations revealed that children high in BI during toddlerhood tended to use a relatively more reactive than proactive control strategy than children low in BI during an AX continuous performance test (AX-CPT) administered at age 13 years (Troller-Renfree et al., 2019). Moreover, the type of cognitive control strategy moderated the relations between BI and parent-reported anxiety such that children with high BI who used a more reactive control strategy had greater total anxiety at age 13 than children with BI who used a more proactive control strategy (Troller-Renfree et al., 2019). Additional support for the idea that children high in both BI and anxiety utilize more in-the-moment type strategies comes from studies showing increased performance on tasks necessitating conflict detection (Thorell et al., 2004; Troller-Renfree et al., 2019; White et al., 2011). Yet, more confirmatory evidence is lent by studies showing increased neural recruitment in high-conflict scenarios (event-related potential; ERPs) by children high in BI. Specifically, relative to youth with BI who are less anxious, youth high in both BI and anxiety have larger P3 responses to novel auditory tones (Reeb-Sutherland et al., 2009), larger N2 responses to conflict (Henderson, 2010; Lamm et al., 2014), and larger error-related negativity (ERN) responses to errors (McDermott et al., 2009), with one study showing that among children with BI, a larger ERN at age 7 prospectively predicted social anxiety symptoms at age 9 (Lahat et al., 2014; but see also Buzzell et al., 2017). The bulk of ERP evidence suggests that children with BI who engage in reactive control more may be at greater risk for anxiety difficulties than children with BI who use a less reactive-like control strategy.

Despite an extant literature focusing on reactive control, no studies examining BI to date have tried to characterize a neural measure of proactive control. This, coupled with the fact that proactive and reactive control processes are thought to be relatively independent (Braver, 2012; Gonthier et al., 2016), leaves it unclear whether the association between BI and anxiety depends purely on the level of reactive control (independent of proactive control) or whether proactive control also plays a critical role in the relations among BI and anxiety. To answer these questions, participants enrolled as part of a longitudinal study were assessed for BI during toddlerhood and completed an AX-CPT task modified for EEG compatibility at age 15 years. Importantly, the present study utilized separate neural measures of proactive and reactive control in order to test whether behavioral findings from an earlier time point of this longitudinal study (Troller-Renfree et al., 2019) were driven mainly by proactive control, by reactive control, or by both. Given the 13-year behavioral finding that BI is associated with anxiety among youth who employ a relatively more reactive than proactive strategy on the AX-CPT (Troller-Renfree et al., 2019), it was hypothesized that, in this new 15-year EEG assessment, BI would be associated with anxiety specifically when proactive control is low and reactive control is high.

## Materials and Methods

### Participants

Participants included 175 adolescents aged 15-17 years who were administered an AX-CPT task during EEG recording as part of a longitudinal study examining the relations between infant temperament and the emergence of anxiety. This study’s recruitment strategy and screening methods have been described in detail elsewhere (Hane et al., 2008). Briefly, 779 infants (age 4 months) completed a laboratory temperament screening for emotional and motor reactivity towards novel auditory and mobile stimuli. From these, infants with high motor and high positive or high negative reactivity were oversampled to reflect a range of temperamental reactivity that is wider than would be found in a randomly selected community sample. The selected infants (n = 291) continued to participate in assessments of cognitive and socio-emotional development throughout childhood and adolescence. Informed consent and assent (as appropriate) were obtained at each assessment, and each visit protocol was approved by the institutional review board of the University of Maryland, College Park.

### Behavioral Inhibition

Replicating the methods used in our previous investigation (Troller-Renfree et al., 2019), BI was assessed at ages 24 and 36 months. Behavioral coding of laboratory assessments (Calkins et al., 1996; Fox et al., 2001) and maternal report of social fear (using the Toddler Behavior Assessment Questionnaire) were standardized and then averaged together to create a BI composite score, based on the assumption that combining data from different informants, contexts, and ages reflects a more comprehensive assessment of the child’s temperament (Lahat et al., 2014; Lamm et al., 2014; Walker et al., 2014).

### AX Continuous Performance Task

To measure distinct neural indices of proactive and reactive control, participants completed an AX-CPT (Barch et al., 1997; Braver, 2012; Cohen et al., 1999) that was modified for simultaneous EEG recording. The AX-CPT presents a continuous series of letter pairs (i.e., a cue letter followed by a probe letter) dissociated into 4 trial types: AX, AY, BX, and BY. AX trials were the target trials, meaning that when participants saw an “A” cue followed by an “X” probe, they were to press a different button in response to the probe than during the other 3 trial types. Specifically, participants were instructed to press “1” following every cue, as well as most probe types, with the exception of an “X” probe that was preceded by an “A” cue; in this case, participants were to press a “4”. Consistent with past ERP studies involving the AX-CPT, the traditional 70%/10%/10%/10% trial breakdown (reflecting AX/AY/BX/BY trials) was modified to 55%/15%/15%/15% in order to increase trial counts necessary to achieve adequate ERP signal-to-noise ratio (Lamm et al., 2013; Troller-Renfree, 2018). Participants completed a total of 319 trials (175/48/48/48) presented in random order. Letter stimuli were presented in boldface 60-point Courier New font on a black background. To make clear the distinction between cues and probes, cues were presented in cyan and probes were presented in white. Each trial began with a center fixation cross, followed by the cue stimulus which was presented for 500 ms. The cue stimulus was followed by a randomized interstimulus interval of 1400-1600 ms (fixation cross). The probe was also presented for 500 ms, with a response window of 1000 ms starting from probe onset. Stimuli were presented on a 17-inch LCD monitor using E-Prime 2.0 Professional (Psychology Software Tools, Inc., Sharpsburg, PA).

Consistent with past work, individual trials were excluded from analysis if reaction time (RT) was >3 standard deviations above or below each participant’s mean RT on correct trials (Troller-Renfree et al., 2019), resulting in exclusion of less than 3% of all trials (*M* = 2.92%, *SD* = 0.01%). After excluding outlier trials, accuracy and mean reaction times were computed for each trial type. Consistent with other studies with children, participants were excluded from analyses if they had less than 60% accuracy on BY trials (n = 2) (Troller-Renfree et al., 2019, 2020), which are the easiest trials because the B cue and Y probe signal totally redundant information. *d’* context, a commonly used behavioral index based on signal detection theory, provides a measure of the ability to discriminate between target and nontarget trials as a function of the cue (Cohen et al., 1999). *d’* context scores were computed by comparing correct responses on AX trials (hits) relative to incorrect responses on BX trials (false alarms). A correction was applied in cases where there was a hit rate of 1 (hit rate = 2^−(1/N)^, where N = number of target trials) or a false alarm rate of 0 (false alarm rate = 1-2^-(1/N)^, where N = number of nontarget trials). The distribution of *d’* context scores (skewness −0.09, kurtosis 2.49) was inspected and determined to be normal. Consistent with the broader literature involving *d’* context, higher scores were interpreted to indicate a more proactive style of cognitive control because the participant used the cue information to inform future responses, whereas lower scores indicate a relatively more reactive style of control because the cue information was less motivationally salient to the participant (Cohen et al., 1999; Troller-Renfree et al., 2019, 2020).

### Screen for Child Anxiety Related Emotional Disorders

Each participant and their parent completed the revised version of the Screen for Child Anxiety Related Emotional Disorders (SCARED) (Monga et al., 2000) at the 15-year assessment. The parent and child versions of the SCARED consist of 41 items presented on a 3-point Likert scale (0 = never/hardly ever true, 1 = sometimes/somewhat true, 2 = very/often true). Total anxiety scores were the primary outcome of interest. In order to combine information from multiple informants while also accounting for differences in how parents and children rate anxiety symptoms (Bowers et al., 2020), total anxiety scores were computed separately for parent and child, Z-transformed, and then averaged together to form an anxiety composite score for analyses (for separate regression results involving child-reported or parent-reported anxiety, see Supplementary Tables S1-S4).

### Electrophysiological Recording, Pre-processing, and Analysis

Continuous EEG was recorded using a 128-channel Geodesic Sensor Net (Electrical Geodesics, Inc., Eugene, OR) and sampled at 250 Hz. Before data collection, all electrode impedances were reduced to < 50 kΩ. During data collection, electrodes were referenced to electrode Cz. All pre-processing, including ocular artifact detection and removal, was performed with the Maryland Analysis of Developmental EEG (MADE) pipeline (Debnath et al., 2020), which utilizes MATLAB (The MathWorks, Natick, MA) functions from EEGLAB (Delorme & Makeig, 2004) and its plugins “FASTER” (Nolan et al., 2010), “ADJUST” (Mognon et al., 2011), and “ADJUSTED ADJUST” (Leach et al., 2020). Offline, data were re-referenced to an average reference and band-pass filtered from 0.3 to 50 Hz with a digital FIR filter. Data were segmented separately for each of the four trial types (i.e, AX, AY, BX, BY). Only trials with correct behavioral responses were analyzed. Channels were marked bad if voltage exceeded ±150 µV, and any epochs in which more than 10% of non-ocular channels exceeded this threshold were marked bad; otherwise, bad channels were interpolated via a spherical-spline interpolation. Participants were only included in subsequent analyses if they had at least 10 artifact-free trials of each of the four trial types (n = 144; *M*_*age*_ = 15.4 years, *SD*_*age*_ = 0.6; 55.9% female). Remaining EEG processing steps were performed with a combination of custom MATLAB scripts and the FieldTrip Toolbox (Oostenveld et al., 2010). A Laplacian transform was applied to convert epoch data from µV to V/m^2^ (i.e., current source density), thus improving spatial resolution (Tenke & Kayser, 2012). In line with previous ERP work involving the AX-CPT, analysis focused on the cue-locked P3b and the probe-locked N2 (van Wouwe et al., 2011). All ERPs were aligned to a baseline of −200 to 0 ms with respect to stimulus onset. Each ERP component was scored by identifying the positive (in the case of the P3b) or negative (in the case of the N2) peak within the scoring window and averaging the amplitude from 40 ms (i.e., 10 sample points) pre-peak to 40 ms post-peak. This adaptive mean scoring approach was used because it is more robust to potential individual differences in peak latency than averaging across the entire scoring window, while still representing an efficient estimation of the true ERP amplitude (Clayson et al., 2013). Sensors for the centroparietal (P3b) and frontocentral (N2) regions-of-interest were selected based on the topography of the grand average waveforms, and the scoring time windows (described below) corresponded to the latencies between which the grand average waveforms exceeded approximately half the peak amplitude (van Wouwe et al., 2011).

### P3b

The cue-locked P3b was used as the measure of proactive control. Previous work has shown that the B-cue minus A-cue P3b difference score significantly mediates the relationship between children’s working memory abilities and their preference for a more proactive (versus reactive) behavioral strategy during the AX-CPT (Troller-Renfree et al., 2020). The P3b was measured within a time window of 430 to 680 ms post-cue after averaging across centroparietal sensor sites (E31, E54, E55, E79, and E80). Analyses focused on the difference between B and A cues (ΔP3b) (Troller-Renfree et al., 2020; van Wouwe et al., 2011).

### N2

The probe-locked N2 was used as the measure of reactive control. A 2-Hz high-pass filter was applied to probe-locked epochs in order to reduce the influence of the probe-locked P3b on the N2 (van Wouwe et al., 2011). The N2 was measured within a time window of 260 to 350 ms post-cue after averaging across frontocentral sensor sites (E5, E6, E7, E12, E13, E106, and E112). In line with the *d’* context behavioral measure of cognitive control strategy, analyses of the probe-locked N2 focused on the difference between AX and BX trials (ΔN2), indicating the extent to which participants discriminated between target and nontarget probes as a function of the cue (Cohen et al., 1999).

### Data Analytic Strategy

To assess whether proactive and reactive control moderated the relations between BI and anxiety, two linear regression models were tested in R (version 3.6.2) with the function “lm”. The outcome variable in both models was parent- and child-reported total anxiety from the SCARED, which, as noted earlier, were Z-transformed and then averaged together to create a composite total anxiety score. The first model was included as a partial replication of our previous investigation (Troller-Renfree et al., 2019). The model included BI, *d’* context (as a behavioral measure of proactive vs. reactive control), and their interaction as predictors. In the second model, predictors included BI, ΔP3b, ΔN2, and their two- and three-way interactions. Outliers were excluded from all between-subjects analyses if they were >3 standard deviations from the sample mean on the variable being tested, and all predictors were mean centered prior to the computation of interaction terms. Simple slopes from interactions were probed with Johnson-Neyman tests (Johnson & Neyman, 1936) with robust standard error estimation using the “sim_slopes” function as part of the R package “interactions” (Long, 2019).

## Results

### Task Behavior

#### Within-Subjects

Descriptive statistics and bivariate correlations for key variables of interest are presented in Table 1. A one-way analysis of variance (ANOVA) revealed a significant within-subjects effect of trial type on correct-trial probe RT (*F*(3, 572) = 254.10, η^2^ = .57, *p* < .001). Post hoc tests revealed that all trial types were different from each other in terms of RT (all *p*s < .05). Specifically, probe RTs were fastest during BX trials (*M* = 287 ms, *SD* = 100 ms), followed by BY (*M* = 293 ms, *SD* = 100 ms), AX (*M* = 340 ms, *SD* = 83 ms), and AY (*M* = 452 ms, *SD* = 95 ms).

**Table 1.** Descriptive Statistics and Pearson Correlations

| Statistic | *N* | *Mean* | *SD* | 1 | 2 | 3 | 4 | 5 | 6 | 7 | 8 | 9 | 10 |
| --- | --- | --- | --- | --- | --- | --- | --- | --- | --- | --- | --- | --- | --- |
| 1. Behavioral Inhibition (Standardized) | 134 | -0.01 | 0.75 |  |  |  |  |  |  |  |  |  |  |
| 2. SCARED Total Anxiety Parent Report | 133 | 10.69 | 9.21 | 0.10 |  |  |  |  |  |  |  |  |  |
| 3. SCARED Total Anxiety Child Report | 125 | 20.48 | 11.75 | 0.03 | 0.54*** |  |  |  |  |  |  |  |  |
| 4. SCARED Total Anxiety Composite (Z-Scored) | 122 | -0.03 | 0.82 | 0.06 | 0.86*** | 0.88*** |  |  |  |  |  |  |  |
| 5. *d'* Context | 142 | 3.14 | 0.62 | -0.08 | 0.14 | 0.04 | 0.10 |  |  |  |  |  |  |
| 6. Cue-Locked P3b A Trials (V/m<sup>2</sup>) | 144 | 1.10E-06 | 6.08E-07 | -0.04 | 0.02 | -0.03 | -0.02 | 0.19* |  |  |  |  |  |
| 7. Cue-Locked P3b B Trials (V/m<sup>2</sup>) | 142 | 1.41E-06 | 7.57E-07 | 0.04 | 0.10 | 0.12 | 0.14 | 0.20* | 0.84*** |  |  |  |  |
| 8. Cue-Locked P3b B-A Difference (V/m<sup>2</sup>) | 141 | 3.23E-07 | 4.02E-07 | 0.06 | 0.08 | 0.23* | 0.23* | 0.14 | 0.09 | 0.61*** |  |  |  |
| 9. Probe-Locked N2 AX Trials (V/m<sup>2</sup>) | 144 | -2.00E-07 | 1.66E-07 | -0.07 | 0.01 | 0.07 | 0.03 | -0.10 | -0.26** | -0.30*** | -0.18* |  |  |
| 10. Probe-Locked N2 BX Trials (V/m<sup>2</sup>) | 142 | -1.46E-07 | 1.49E-07 | -0.09 | 0.12 | 0.12 | 0.14 | -0.21* | -0.20* | -0.24** | -0.10 | 0.38*** |  |
| 11. Probe-Locked N2 AX-BX Difference (V/m<sup>2</sup>) | 141 | -4.38E-08 | 1.65E-07 | 0.00 | -0.04 | -0.01 | -0.05 | 0.08 | -0.01 | -0.02 | -0.11 | 0.60*** | -0.48*** |

Note. All values reflect exclusion of outliers (see Data Analytic Strategy). \* $p < .05$ . \*\* $p < .01$ . \*\*\* $p < .001$ .

A similar one-way ANOVA revealed a significant within-subject effect of trial type on accuracy (*F*(3, 572) = 170, η^2^ = .47, *p* < .001). Post hoc tests revealed that all trial types were different from each other (all *p*s < .05). BY trials were most accurate (*M* = 96.9%, *SD* = 4.4%), followed by BX (*M* = 93.4%, *SD* = 7.9%), AX (*M* = 92.0%, *SD* = 5.3%), and AY (*M* = 75.7%, *SD* = 13.8%). Consistent with past studies of the AX-CPT in populations relying on proactive control, AY trials were the slowest and least accurate of all the trial types.

#### Between-Subjects

The results of the regression model involving *d’* context are presented in Table 2. There were no significant main effects of BI or *d’* context and no significant interaction effect (all *p*s > .05).

**Table 2.** d’ Context Regression Model Predicting Total Anxiety (Z-Scored)

| *Predictors* | *Standardized*<br>*Beta* | *95% CI* | *p* |
| --- | --- | --- | --- |
| (Intercept) | 0.012 | -0.171 – 0.195 | 0.897 |
| Behavioral Inhibition (BI) | 0.043 | -0.142 – 0.228 | 0.644 |
| *d'* Context | 0.111 | -0.076 – 0.297 | 0.242 |
| BI * *d'* Context interaction | 0.099 | -0.084 – 0.281 | 0.286 |
| Observations | 119 |  |  |
| R<sup>2</sup> / R<sup>2</sup> adjusted | 0.021 / -0.005 |  |  |

### EEG

#### Within-Subjects

For grand average cue- and probe-locked ERPs, see Figure 1. Cue-locked P3B amplitude was significantly more positive following B cues than following A cues (*t*(143) = 9.51, *d* = 0.79, *p* < .001). A one-way ANOVA revealed a significant within-subjects effect of trial type on probe-locked N2 amplitude (*F*(3, 572) = 44.38, η^2^ = .19, *p* < .001). Post hoc tests revealed that all trial types significantly differed from each other in terms of N2 amplitude (*p*s < .01) with the exception of AX and BY (*p* = .654).

**Figure 1.**
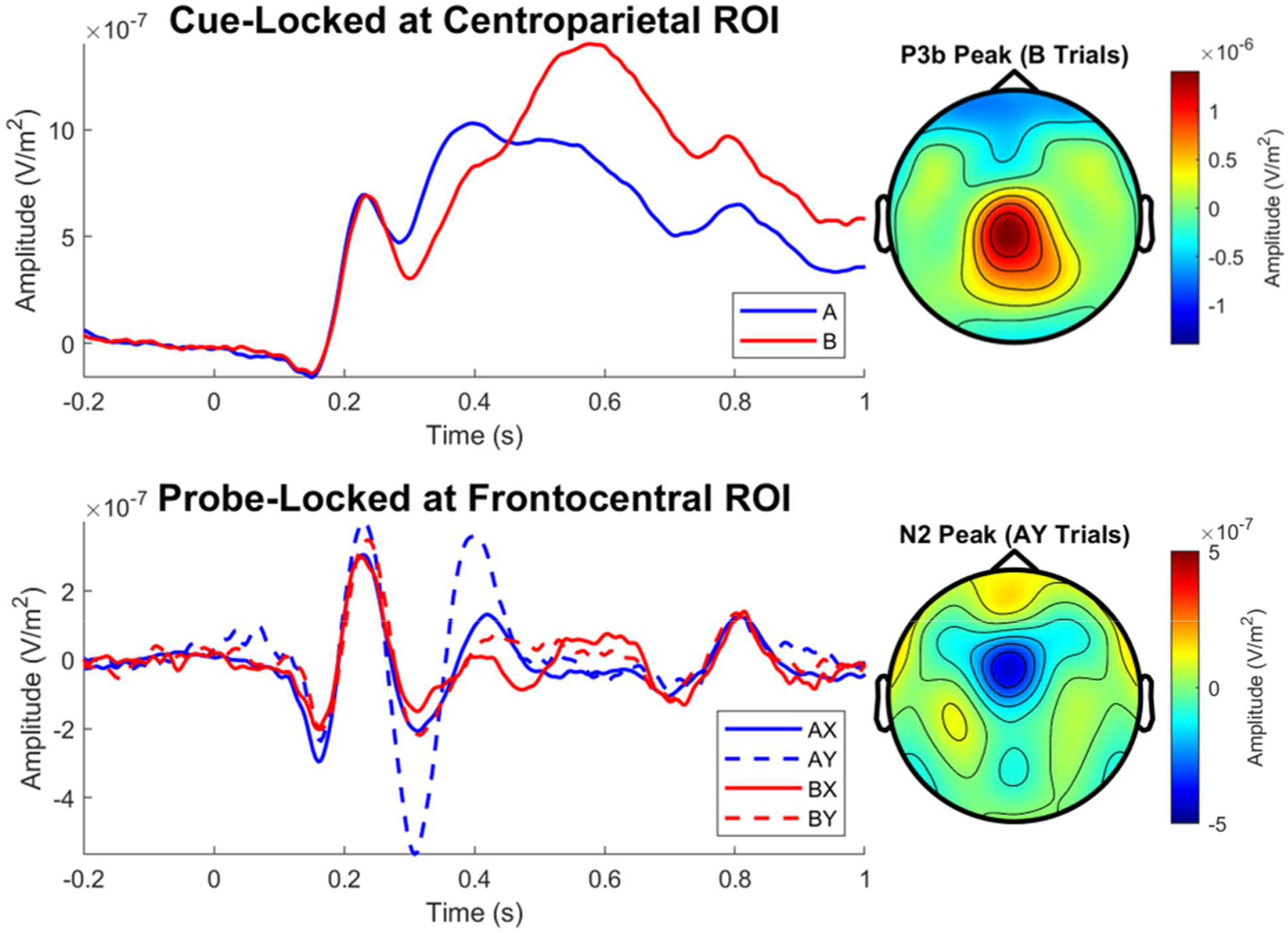
Grand Average ERP Waveforms. *Note*. Centroparietal region-of-interest (ROI) included electrodes E31, E54, E55, E79, and E80. Frontocentral ROI included electrodes E5, E6, E7, E12, E13, E106, and E112. The cue-locked P3b was significantly more positive following B cues than following A cues (*p* < .001). With regard to the probe-locked N2, all trial types significantly differed from each other (*p*s < .01) with the exception of AX and BY (*p* = .654).

#### Between-Subjects

The results of the regression model involving EEG measures are presented in Table 3. There was a significant main effect of ΔP3b such that a larger B-minus A-cue difference (indicating greater use of a proactive strategy) was associated with greater anxiety (*β* = .283, *p* = .003). There was also a significant three-way interaction between BI, ΔP3b, and ΔN2 (*β* = .232, *p* = .017; see Figure 2, panel A). A Johnson-Neyman follow-up test revealed that BI is significantly associated with greater anxiety (*p* < .05) specifically when ΔP3b is small (indicating a less proactive strategy; ΔP3b Z < 1) and ΔN2 is large (i.e., more negative, indicating a more reactive strategy; ΔN2 Z < -.75; see Figure 2, panel B).

**Table 3.** ERP Regression Model Predicting Total Anxiety (Z-Scored)

| *Predictors* | *Standardized*<br>*Beta* | *95% CI* | *p* |
| --- | --- | --- | --- |
| (Intercept) | -0.007 | -0.185 – 0.171 | 0.935 |
| Behavioral Inhibition (BI) | 0.059 | -0.121 – 0.239 | 0.515 |
| $\Delta N2$ | 0.011 | -0.188 – 0.210 | 0.911 |
| $\Delta P3b$ | 0.283 | 0.099 – 0.467 | 0.003 |
| BI * $\Delta N2$ interaction | -0.117 | -0.325 – 0.091 | 0.267 |
| BI * $\Delta P3b$ interaction | -0.034 | -0.227 – 0.159 | 0.731 |
| $\Delta N2$ * $\Delta P3b$ interaction | -0.097 | -0.284 – 0.089 | 0.303 |
| BI * $\Delta N2$ * $\Delta P3b$ interaction | 0.232 | 0.041 – 0.422 | 0.017 |
| Observations | 116 |  |  |
| R<sup>2</sup> / R<sup>2</sup> adjusted | 0.129 / 0.072 |  |  |

**Figure 2.**
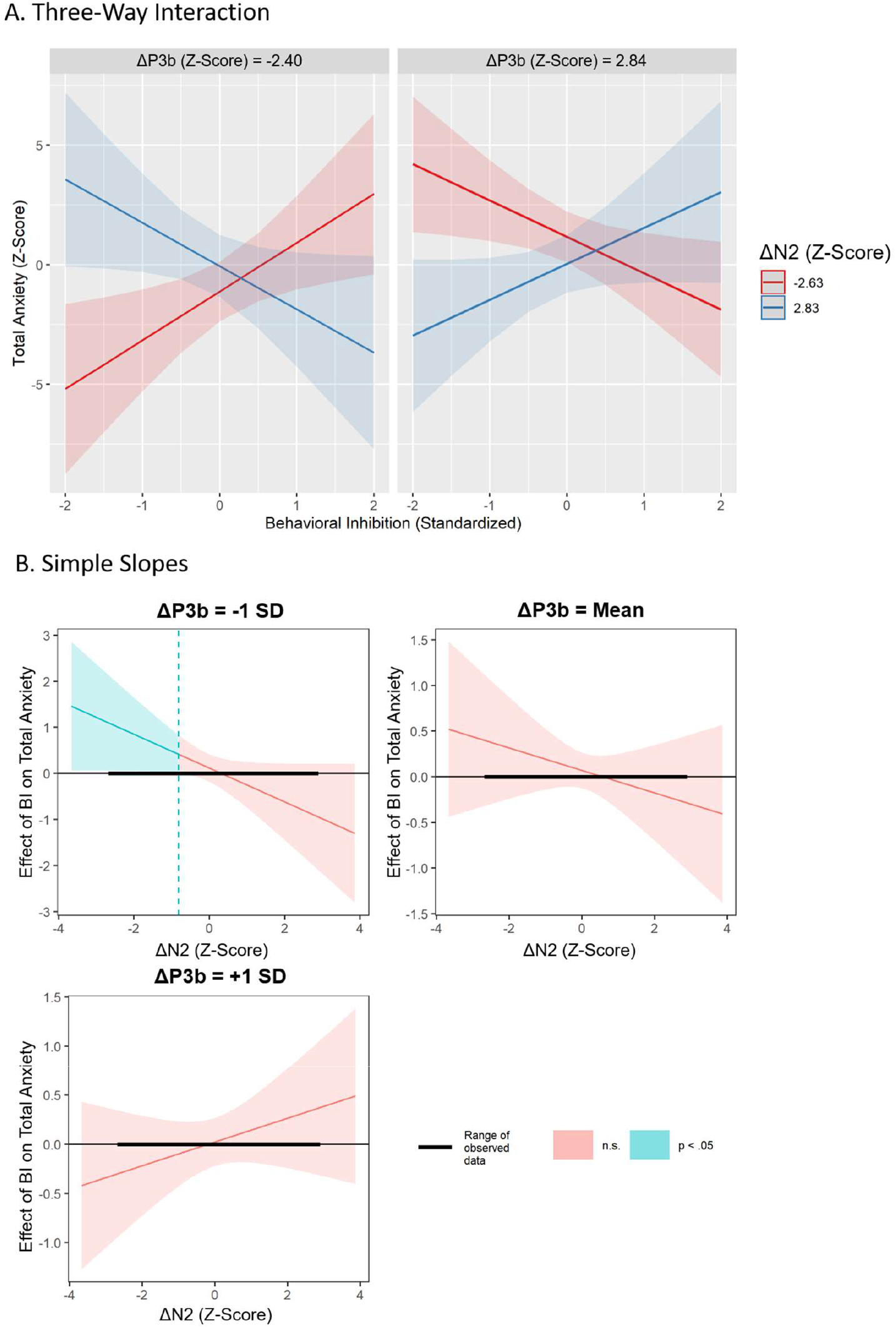
Three-Way Interaction and Simple Slopes. *Note*. A) The three-way interaction between BI, ΔP3b, and ΔN2 (*p*_interaction_ = .017), and B) Johnson-Neyman plots illustrating results of simple slopes analysis, which revealed that BI is significantly associated with greater anxiety (*p* < .05) when ΔP3b is small (i.e., less positive; ΔP3b cutoff: Z < 1) and ΔN2 is large (i.e., more negative; ΔN2 cutoff: Z < -.75).

## Discussion

The present study tested whether the relations between early BI and adolescent anxiety vary as a function of reactive and proactive control. In light of previous 13-year behavioral findings from this sample indicating that a greater relative reliance on reactive control (compared to proactive control) predicted risk for anxiety among children high in BI (Troller-Renfree et al., 2019), we hypothesized that early BI would be associated with greater anxiety specifically among adolescents using a strategy characterized by low proactive control and high reactive control at age 15 years. Because the behavioral measure of proactive versus reactive control (i.e., *d’* context) is essentially a difference score indicating a participant’s relative preference for one strategy versus the other, it was important to use separate neural measures for proactive and reactive control in order to test whether past behavioral findings were driven mainly by proactive control, by reactive control, or by both. Using the cue-locked ΔP3b as a measure of proactive control and the probe-locked ΔN2 as a measure of reactive control, we found that the relations between early BI and greater anxiety are only significant among adolescents who use a cognitive control strategy characterized by low proactive control (i.e., ΔP3b is smaller/less positive) and high reactive control (i.e., ΔN2 is larger/more negative). However, we did not replicate the 13-year behavioral findings at this 15-year assessment, possibly due to differences in task parameters or due to developmental changes in cognitive control and anxiety. Nevertheless, ERP results indicated that both proactive and reactive control processes influence the relations amongst BI and anxiety.

The present findings support an emerging view of BI’s neurophysiological profile (Buzzell et al., 2018; Fox et al., under review; Henderson et al., 2015; Henderson & Wilson, 2017). According to this view, although BI is associated with heightened detection of salient stimuli (e.g., threatening faces, novel auditory tones), some children with BI learn to moderate their responses to novelty or unfamiliarity over time via increased proactive control. This increased proactive control helps the child recover their goal-oriented attention and reduces the length of time that attention is shifted toward a salient stimulus when it occurs, thereby reducing risk for anxiety. In contrast to proactive control, reactive control *maintains* attention toward the salient stimulus (thus, increasing risk for anxiety) in order to resolve conflict or support quick and reflexive corrections to behavior. Together, these two types of control influence the child’s ability to fluidly respond to salient stimuli in goal-directed contexts; yet, they have distinguishable associations with anxiety.

In addition to the BI pathway described above, results revealed that adolescents with a more positive ΔP3b, which we interpreted as indicating a highly proactive strategy, tended to have greater total anxiety. Importantly, this ΔP3b effect was independent of BI and there was no significant ΔP3b * BI interaction, indicating that greater reliance on proactive control may reflect a general risk pathway for anxiety. This raises the question, however, of how proactive control can be a risk factor for anxiety while also being protective against anxiety among youth high in BI. One possibility is that whereas a certain minimum level of proactive control may be needed for children with BI to avoid being unduly disrupted by salient stimuli, an excessive, over-reliance on proactive control might reduce children’s ability to flexibly respond to salient events as they arise, regardless of BI status. In other words, proactive control may be protective when salience detection is elevated (as in the case of BI), but instead increases risk for anxiety when salience detection is at lower levels. In order to test this hypothesis, future work would benefit from the inclusion of a salience detection measure (e.g., a measure of threat bias from a dot-probe task) in addition to measures of proactive and reactive control.

Another possibility is that the etiological pathway to anxiety associated with proactive control and the pathway to anxiety associated with BI each give rise to a different subtype of anxiety. Although BI is a strong risk factor for the development of future anxiety (Chronis-Tuscano et al., 2009; Clauss & Blackford, 2012), it is important to note that many children without a history of BI also go on to develop anxiety difficulties later in life. Thus, there may be multiple pathways through which a child can develop problems with anxiety, and it could be the case that each pathway/subtype is associated with a different profile of cognitive control dynamics. Previous neuroimaging work has demonstrated that cognitive-control-related neural activity differentiates BI and non-BI anxious youth (Abend et al., 2020; Smith et al., 2019), suggesting that cognitive control could serve as a potential criterion for defining anxious subtypes. However, the present study is the first to characterize how proactive and reactive control each uniquely relate to these pathways/subtypes.

In addition to having implications for the conceptualization of heterogeneous anxiety, the present findings may also inform anxiety intervention efforts. If it is the case that proactive control protects youth with BI against anxiety, proactive control may be a promising intervention target for this subgroup of children. Although interventions targeting salience detection (e.g., attention bias modification) (MacLeod & Mathews, 2012) are likely appropriate for all anxious youth, children with BI may respond better to an intervention aiming instead to enhance proactive control, as heightened salience detection is a core feature of BI (Fox et al., 2005; Kagan et al., 1984) and thus may be less malleable for this group than is proactive control.

Critically, the present study does not allow for causal inferences in the associations between cognitive control processes and anxiety, in part due to the lack of temporal separation (i.e., with the exception of BI, which was assessed during toddlerhood, all other measures were assessed at the same 15-year time point) but also due to the lack of random assignment. Moreover, the present longitudinal sample was oversampled for extreme levels of motor and positive or negative reactivity during toddlerhood (Hane et al., 2008), possibly limiting generalizability; however, this oversampling approach was necessary in order to achieve a sufficient number of children with BI, which is seen in approximately 10-15% of young children (Fox et al., 2005). Nevertheless, in addition to benefitting from early laboratory assessment of early BI (as opposed to relying on retrospective report), the present study was aided by a relatively large sample size which provided sufficient statistical power to detect the interaction between BI, proactive control, and reactive control.

In summary, the present findings suggest that early BI is associated with elevated anxiety symptoms among adolescents who rely more on reactive control strategies (as indicated by a larger ΔN2) and less on proactive strategies (as indicated by a smaller ΔP3b), possibly indicating that, among children with early BI, proactive control is protective against anxiety whereas reactive control increases anxiety risk. In addition, results revealed a non-BI pathway to anxiety involving elevated proactive control, suggesting that proactive control distinguishes pathways to anxiety occurring only with BI from that occurring regardless of BI level.

## Data Availability

The data that support the findings of this study may be made available from the corresponding author, E. A. V., upon reasonable request.

## Acknowledgements

This research was supported by the National Institute of Mental Health, Grand/Award Number: U01MH093349 (to N. A. F.).

## Key Points

- Behaviorally inhibited (BI) temperament is a strong predictor of anxiety problems later in life, but this association is moderated by cognitive control factors.
- By separating proactive and reactive control processes using EEG, the present study is the first to characterize how proactive and reactive control uniquely relate to pathways toward anxiety risk.
- Findings suggest that BI relates to risk for anxiety specifically among adolescents who rely less on proactive strategies and more on reactive control strategies.
- Results also revealed that proactive control differentiates a BI-related etiological pathway to anxiety from a more general pathway to anxiety occurring regardless of BI level.
- Overall, findings indicate that cognitive control could serve as a potential criterion for defining anxious subtypes.

