## Supplementary Tables S1-S4 for "Behavioral Inhibition and Dual Mechanisms of Anxiety Risk: Disentangling Neural Correlates of Proactive and Reactive Control"

Supplementary Table S1

*d’ Context Regression Model Predicting Parent-Reported Total Anxiety (Z-Scored)*

| *Predictors* | *Standardized Beta* | *95% CI* | *p* |
| --- | --- | --- | --- |
| (Intercept) | 0.005 | -0.169 – 0.178 | 0.472 |
| Behavioral Inhibition (BI) | 0.088 | -0.088 – 0.264 | 0.341 |
| *d'* Context | 0.147 | -0.028 – 0.322 | 0.101 |
| BI * *d'* Context interaction | 0.062 | -0.109 – 0.233 | 0.475 |
| Observations | 130 |  |  |
| R^2^ / R^2^ adjusted | 0.031 / 0.008 | |  |

Supplementary Table S2

*d’ Context Regression Model Predicting Child-Reported Total Anxiety (Z-Scored)*

| *Predictors* | *Standardized Beta* | *95% CI* | *p* |
| --- | --- | --- | --- |
| (Intercept) | 0.010 | -0.173 – 0.192 | 0.968 |
| Behavioral Inhibition (BI) | 0.017 | -0.168 – 0.202 | 0.890 |
| *d'* Context | 0.062 | -0.123 – 0.248 | 0.514 |
| BI * *d'* Context interaction | 0.084 | -0.099 – 0.268 | 0.365 |
| Observations | 121 |  |  |
| R^2^ / R^2^ adjusted | 0.010 / -0.016 | |  |

Supplementary Table S3

*ERP Regression Model Predicting Parent-Reported Total Anxiety (Z-Scored)*

| *Predictors* | *Standardized Beta* | *95% CI* | *p* |
| --- | --- | --- | --- |
| (Intercept) | -0.008 | -0.183 – 0.166 | 0.441 |
| Behavioral Inhibition (BI) | 0.114 | -0.063 – 0.290 | 0.202 |
| ΔN2 | 0.020 | -0.174 – 0.214 | 0.816 |
| ΔP3b | 0.080 | -0.100 – 0.259 | 0.319 |
| BI * ΔN2 interaction | 0.027 | -0.174 – 0.228 | 0.797 |
| BI * ΔP3b interaction | -0.140 | -0.325 – 0.046 | 0.127 |
| ΔN2 * ΔP3b interaction | -0.198 | -0.378 – -0.018 | 0.031 |
| BI * ΔN2 * ΔP3b interaction | 0.061 | -0.119 – 0.241 | 0.504 |
| Observations | 127 |  |  |
| R^2^ / R^2^ adjusted | 0.081 / 0.027 | |  |

Supplementary Table S4

*ERP Regression Model Predicting Child-Reported Total Anxiety (Z-Scored)*

| *Predictors* | *Standardized Beta* | *95% CI* | *p* |
| --- | --- | --- | --- |
| (Intercept) | -0.002 | -0.181 – 0.176 | 0.836 |
| Behavioral Inhibition (BI) | 0.037 | -0.144 – 0.217 | 0.635 |
| ΔN2 | 0.058 | -0.141 – 0.257 | 0.558 |
| ΔP3b | 0.283 | 0.100 – 0.467 | 0.002 |
| BI * ΔN2 interaction | -0.121 | -0.329 – 0.086 | 0.242 |
| BI * ΔP3b interaction | -0.052 | -0.243 – 0.138 | 0.518 |
| ΔN2 * ΔP3b interaction | -0.060 | -0.245 – 0.125 | 0.518 |
| BI * ΔN2 * ΔP3b interaction | 0.195 | 0.008 – 0.382 | 0.041 |
| Observations | 118 |  |  |
| R^2^ / R^2^ adjusted | 0.107 / 0.050 | |  |
